# Arm choice post-stroke is habitual rather than optimal in right-, but not in left-paretic individuals

**DOI:** 10.1101/2020.08.31.20185389

**Authors:** Sujin Kim, Cheol E. Han, Bokkyu Kim, Carolee J. Winstein, Nicolas Schweighofer

## Abstract

In non-disabled individuals, arm choice in pointing movements depends on expected biomechanical effort, expected success, and a handedness bias. Following a stroke, is arm choice re-optimized to account for the decreased motor performance, or does it follow a pre-injury habitual pattern? Because premorbidly right-handed individuals with left hemiparesis generally use their affected arm less than those with right hemiparesis, we hypothesized that arm choice follows a more habitual pattern in right-than in left-hemiparetic individuals. Participants with mild to moderate chronic stroke who were right-handed before stroke performed pointing movements in both free- and forced-choice blocks, both under a no-time constraint condition and under a fast-time constraint condition designed to promote habitual choice. Mixed-effects models of arm choice revealed that expected effort and side of stroke predicted choices overall. However, expected success predicted choice in left-, but not of right-hemiparetic individuals. Furthermore, while left-hemiparetic individuals tended to avoid unsuccessful movements in the fast condition by selecting their non-paretic arm, right-hemiparetic individuals persevered in choosing their more affected arm. In addition, reaction times decreased in left-hemiparetic individuals between the no-time and the fast condition but showed no changes in right-hemiparetic individuals. Finally, arm choice in the no-time condition correlated with a clinical measure of spontaneous arm use for right-, but not for left-hemiparetic individuals. Our results thus show that, in premorbidly right-handed individuals with mild to moderate chronic stroke, arm choice is habitual in right-hemiparetic individuals, but shows a greater degree of optimality by taking account expected success in left-hemiparetic individuals.

New & Noteworthy

Although we are seldom aware of it, we constantly make decisions to use one arm or the other in our daily activities. Here, we study whether these decisions change following a chronic mild to moderate stroke that affects motor control. Our results show that chronic stroke survivors with a right hemiparesis make arm choice using a habitual strategy, while those with left hemiparesis re-optimize their choices to account for their impaired motor performance.

## Introduction

Consider the apparently simple task of reaching a target as fast as possible. What arm should one choose? In a previous study of arm choice for horizontal movements in right-handed non-disabled subjects (Schweighofer et al. 2015), we showed that, for a given target, the arm that maximizes expected success and minimizes expected effort is chosen with higher probability than the other arm, with an overall handedness bias favoring the right arm. How does this pattern of arm choice change in chronic stroke survivors who experience mostly unilateral motor deficits? In particular, is arm choice re-optimized to account for the decreased performance in the more affected arm, or does it follow a pre-injury habitual pattern?

To address this question, we adopted a contemporary decision framework according to which human choices are driven by a combination of a goal-oriented system and a habitual system (Kable and Glimcher 2009; Daw et al. 2011). The goal-directed system makes choices via mental simulation of the decision environment to evaluate the outcomes of possible actions. Thus, decisions governed by the goal-directed system can be quickly modified following environment changes to maintain optimal actions. However, the mental simulations requires relatively long reaction times for decisions (Keramati et al. 2011). In contrast, the habitual system performs choices through a direct comparison among learned expected outcomes (or “values”) for each potential action (Daw et al. 2005; Gläscher et al. 2010). Due to its relative inflexibility, the habitual system generates sub-optimal behavior in response to environmental changes, such as reward devaluation, by maintaining pre-change patterns. However, since the habitual system simply retrieves the values from memory without mental simulation, it is fast, e.g., with short reaction times (Daw et al. 2005). As a result, decision making under time-pressure can enhance the expression of the time-insensitive habitual system (Keramati et al. 2011; Hardwick et al. 2019). Here, we therefore compare arm choices made by stroke survivors with mild to moderate impairments in a no-time constraint condition to fast condition, but not the no-time condition, show good test-retest reliability and correlate with a measure of spontaneous arm use in individuals with chronic stroke hemiparesis (Kim et al. 2018).

In addition to expected success and expected effort, the side of hemiparesis also needs to be considered as a factor in arm choice post-stroke. Previous research demonstrates that while overall use of the paretic arm decreases after stroke in premorbidly right-handed individuals, right hemiparetic individuals (RH) use their more-affected arm to a greater extent than left hemiparetic (LH) individuals (Haaland et al. 2012; Mani et al. 2013, 2014; Bailey et al. 2015). In addition, motor control strategies are different for the dominant and non-dominant arms (Sainburg and Schaefer 2004; Haaland et al. 2012). In particular, in non-disabled right-handed individuals, the right, arm, which is primarily under the control by the left hemisphere, is better than the left at producing precise movement trajectories through feedforward control (Coelho et al. 2013), and is primarily used as the main mover in daily activities (Vega-González and Granat 2005; Michielsen et al. 2012). Indeed, in previous work in pointing movements with right-handed participants, we showed that even when the biomechanical effort is highly favorable to the left arm, there is an overall bias towards using the right arm, with this bias increasing when higher precision is required (Schweighofer et al. 2015).

We therefore hypothesized that, after stroke in premorbidly right-dominant individuals, a habitual pattern of arm choice for reaching movements will be maintained in RH individuals even after repeated unsuccessful movements (which are akin to reward devaluation) due to decreased performance of the more affected arm. In contrast, because left arm use in reaching movements was less habitual pre-stroke, LH individuals will exhibit goal-directed, more optimal, use of their more affected arm. Specifically, if arm choice is habitual, participants will not change their arm choice nor their reaction times when success with the paretic arm is reduced in the fast-time constraint. In contrast, if arm choice is goal-directed, both paretic arm choices and reaction times will decrease in the fast-time condition compared to the non-time condition.

To test the sensitivity of choice to expected success in both RH and LH participants, we analyzed target-specific choices with a mixed-effect logistic regression model of arm choice that extends our previous model in non-disabled individuals (Schweighofer et al. 2015). Consistent with this previous study, we found here that expected biomechanical effort largely influences arm choice in post-stroke individuals. However, we found that expected success differently influenced arm choice between the RH and the LH groups: as LH individuals were sensitive to failures, they were more likely to switch their arm choice from the paretic to the non-paretic side in the fast time constraint condition. In contrast, RH individuals were less likely to switch arm regardless of the time constraints. In addition, reaction times decreased in left-hemiparetic individuals between the no-time and the fast condition, but showed no changes in right-hemiparetic individuals. Finally, we found that arm choice in the no-time constraint condition correlates with a clinical measure of habitual arm use, the Actual Amount of Use Test (AAUT), in right-, but not left-hemiparetic individuals.

## Methods and Methods

### Participants

Data from twenty-two individuals post-stroke with mild to moderate impairment, twelve with RH (1 female, means ± SD aged 62.83 ± 13.95 years) and ten with LH (4 females, mean ± SD aged 56.90 ± 13.53 years), were analyzed. The participants had had their stroke from 0.47 years to 14.38 years before participation. In addition, eleven age-matched nondisabled participants (Controls; 6 females, mean ± SD aged 54.36 ± 10.58 years) were recruited for comparison.

Participants post-stroke were a sub-set of those included in the DOSE rehabilitation clinical trial, for whom arm choice data were available. The DOSE trial was designed to investigate the effect of therapy dose on arm/hand function (NCT 01749358; Winstein et al. 2019). Here, we only analyzed arm choice and performance data at “baseline” before any therapy intervention. Detailed inclusion criteria for participants were previously reported (Kim et al. 2018). Notably, participants post-stroke were included if they had a stroke more than 5 months ago; had mild to moderate motor impairment (upper extremity Fugl-Meyer motor (UEFM) > 19 out of 66); could reach with the paretic arm from the home position to a target 25 cm away within 5 seconds; had no arm/hand neglect as determined by Albert Test; and self-reported premorbidly right-handed. Control participants were included if they reported no prior neurological disorders and self-reported to be right-handed. The study was approved by the Human Research and Review Committee of the University of Southern California and each participant signed and received a copy of an informed consent. Clinical assessments for screening, UEFM and Amount of Arm Use Test (AAUT), as well as arm choice tests were performed by two trained and standardized experimenters.

### Experimental setup and design

#### Experimental Setup

Details of the experimental set-up, shown in Figure 1A, can be found in (Kim et al. 2018). Briefly, participants sat in a wooden chair in front of a table with a restraining belt to minimize compensatory trunk motion during pointing movements. At each trial, a target, appearing at one of the pre-defined 35 target locations, was projected on the table from an overhead projector. Participants were instructed to move their index finger from the home position to the target as rapidly and accurately as possible and to keep their finger on the target for at least 0.5 s before returning to the home position. A pleasant sound was provided following successful reaches, i.e., when the target was reached within the given time; an unpleasant sound was provided otherwise. In addition, participants received a “reward” point for each successful trial. Two Mini-Bird magnetic sensors (Ascension Technology Corporation) were placed on the nail of the right and the left index fingers to measure hand trajectory and arm choice (sampling 100 Hz). Position data from the magnetic sensors were filtered at 5Hz with a Butterworth low pass filter.

**Figure 1.**
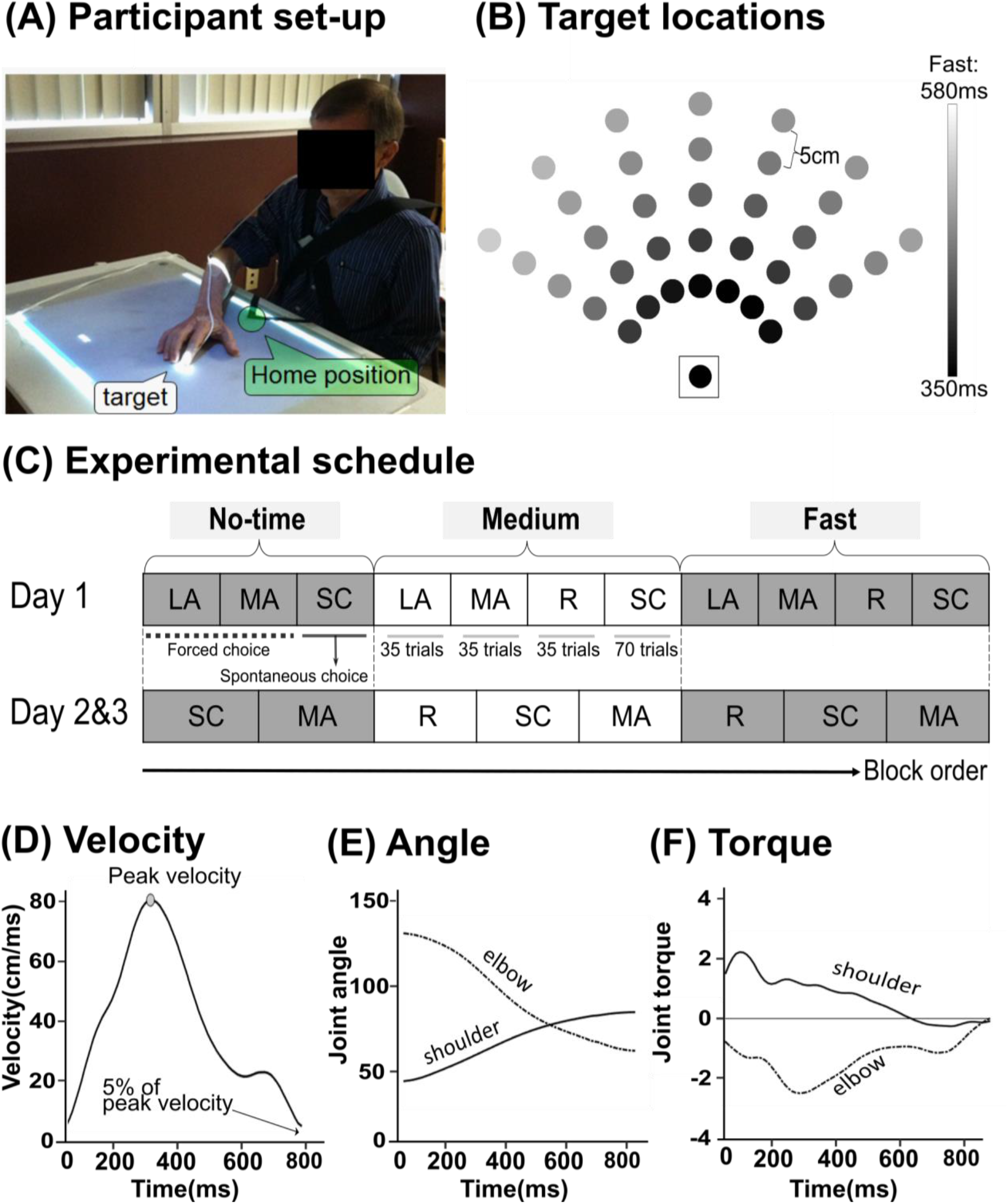
Bilateral Arm Reaching Test (BART) set up, target locations, and protocol. A. Set-up: The green circle and the white circle show the home position and a target, respectively. At each trial, participants were instructed to reach to the target using either right or left index finger (depending on conditions) as quickly and accurately as possible. Magnetic sensors were attached to the index finger tips of both arms to record choice of hand and kinematics. B. The 35 targets displayed in the BART workspace (plus the home target, shown surrounded by a square). Gray shading shows the time constraint for each target in the fast condition. C. Experimental protocol common for the stroke group. In Day1, the forced choice blocks (LA and MA) were presented before the spontaneous choice blocks (SC) for the familiarization purpose. In Days 2 and 3, reminder conditions were presented before the spontaneous choice blocks, followed by the forced choice blocks (MA) for the medium and fast conditions (see Methods). The no-time and fast time constraint conditions were used for the analysis and were shaded by gray color. D. Examples of velocity profile of a single reaching movements for one subject post-stroke illustrating the computation of movement time. E. Example of estimated joint angle. An inverse kinematics model was used to estimate shoulder and elbow joint angles from hand trajectories. F. Example of shoulder and elbow joint toques estimated by an inverse dynamics model of the arm in 2 dimensions. No-time, no-time constraint condition; Medium, medium time constraint condition; Fast, fast time constraint condition; MA, more-affected arm only block; LA, less-affected arm only block; SC, spontaneous choice block; R, reminder block.

In the no-time constraint condition, the targets were displayed until the participants’ index finger landed inside the target disk; in contrast, in the fast-time constraint condition, the targets disappeared ~0.5 sec after movement onset. Movement time limits in the fast-time constraint condition were estimated from previous aiming data in non-disabled right-handed participants (Park et al. 2015), and ranged between 350 ms and 580 ms (see Figure 1B).

#### Experimental Design

Participants performed blocks of forced choice trials and spontaneous choice trials, in no-, medium-, and fast-time constraint conditions to an array of 35 targets (see Figure 1B). In the forced choice blocks participants reached each of the 35 targets once with each arm. In the spontaneous choice blocks participants reached to each target with the arm of their choice, with two trials per target (thus for a total 70 targets, see Figure 1C). The no-time constraint condition was given before the fast-time constraint condition (Figure 1C), because we observed in a pilot study that stroke participants often did not select their more-affected arm when they started with the more challenging condition first. Similarly, the medium-time constraint was included because we noticed in a second pilot study that some participants stopped using their more-affected arm entirely if the fast-time constraint condition directly followed the no-time constraint condition. For simplification, we do not report here choices in the medium-time constraint condition. Reminder sessions, which were similar to the spontaneous choice blocks but with fewer trials (35 trials), were included before the medium and fast-time constraint conditions to remind the participants of the time limits for these condition (See Figure 1C).

#### Stroke participants

Participants post-stroke were tested with the arm-reaching task on Days 1, 2, and 3, with at least 2 weeks between testing days to minimize learning effects on the test. In the forced choice blocks on Day 1, participants reached all 35 targets first with their less-affected arm and then with their paretic, more-affected, arm. In the forced choice blocks on Day 2 and 3, participants reached all targets with their paretic arm only. In the spontaneous choice block in the fast-time constraint condition, participants were instructed to minimize unsuccessful trials either by moving fast or by using the less-affected arm when expecting failure with the more-affected arm. We previously demonstrated excellent test-retest reliability of arm choice in the spontaneous choice block of the fast-time constraint condition and good reliability in the spontaneous choice block of the no-time constraint condition in the same stroke participants (Kim et al. 2018).

#### Control participants

Control participants were tested on Days 1 and 2, with 1 week between testing days. In forced choice blocks on Day 1, control participants reached to all 35 targets first with their right arm and then with their left arm, while on Day 2 they reached the targets with their right arm only. Spontaneous choice blocks were given on Days 1 and 2.

### Data measurement and processing

#### Overall probability of arm choice

Probability of arm choice for each target in each condition was measured from spontaneous choice blocks on Days 1, 2 and 3 for the stroke group and on Days 1 and 2 for the control group (Figure 1C).

#### Overall success rate

Success rates for the more-affected arm in each condition were computed from forced choice blocks on Days 1, 2 and 3 for the stroke group, and on Days 1 and 2 for the control group.

#### Effort estimation

Effort for both arms was estimated from movement trajectories in the forced choice block on Day 1. We assumed planar movements, and performed inverse kinematics followed by an inverse dynamics transformation with left and right planar 2DOF arm models that include a shoulder and an elbow joint, as in our previous study (Schweighofer et al. 2015). The index finger position (x,y) was first transformed into joint angles (i.e., shoulder and elbow joint angles) using inverse kinematics of a 2 DOF arm model (Figure 1E). The shoulder and elbow torques were then estimated using inverse dynamics (Figure 1F). All arm parameters were taken from (Van Beers et al. 2004). Note that we computed “absolute effort” defined by summing the absolute torques at both joints from the movement start to the end, since such effort has been recently shown to better account for arm movement planning (Shadmehr et al. 2016) and was a better predictor of choice than effort derived from summing the squared torques.

#### Reaction times

Reaction times were computed in each spontaneous choice block in both time constraint conditions. Reaction time was defined as the time between the target appearance and the time at which the index finger’s tangential velocity exceeded 5% of maximum velocity (Figure 1D).

### Clinical assessment of arm use

We administered the Actual Amount of Use Test (AAUT) to the stroke participants to assess spontaneous use of the paretic arm and hand in the real-world (Taub et al. 1998). In the AAUT, participants perform 14 upper-extremity daily tasks, such as opening a file folder, and writing on and folding up a piece of paper with the hands of their choice (including bimanually) without any prompts. A trained evaluator watched video recordings of the test and graded spontaneous arm use behaviors based on quality of movement scale (QOM). The QOM score for each item was averaged over the 14 tasks. If participants habitually use their more-affected arm, then we expect arm choice measured by our reaching system should correlate with arm use as assessed by the covert AAUT.

### Overall behavioral and clinical measures analyses

To investigate the effects of time constraint on all behavioral data including choice of paretic arm, effort, success rate, and reaction time, we developed linear mixed-effect models in which condition (no-time and fast-time constraints) and side of stroke (RH and LH) were entered as fixed effects and participants as random intercepts. Log-likelihood ratio tests (LRT) were used to verify the need for inclusion of the random effect term. Visual inspection of residual versus fit plots and qq-plots was performed to check for heteroscedasticity or non-normal distribution of the residuals. When needed, post-hoc analyses were performed using the Tukey test to correct for multiple comparisons. In addition, correlations between the arm choice measured by the reaching system and arm use measured by AAUT in both stroke groups (LH and RH) were conducted using spearman’s correlation after Shapiro-Wilk test for normality. Finally, between-group differences in stroke onset and UEFM were analyzed using either independent t-test or Wilcoxon rank sum test after Shapiro-Wilk test for normality. Possible differences in age for RH, LH, and Control groups were tested using the Kruskal-Wallis test. All statistical analyses were performed with the R statistical package version 3.6.2 (R Core Team 2012). Significance levels were set to p = .05 for these overall analyses and all results are given in mean ± standard error.

### Target-by-target arm choice analysis: Mixed-effect logistic regression models

To test whether the probability of arm choice measured in the spontaneous choice blocks was different between RH and LH groups, and whether that probability is predicted by expected success and expected effort measured in the forced choice blocks, we developed mixed-effect logistic regression models of arm choice. The models are based on our previous model for non-disabled individuals, in which we showed that between-arm differences in expected effort and expected success accurately predicted arm choice (Schweighofer et al. 2015). Here, we tested the following regressors for inclusion in the stroke model: 1) between-arm differences in effort, 2) between-arm differences in success, and 3) side affected by stroke, which was entered as a binary variable (0 for RH and 1 for LH). To take into account large differences in participants’ characteristics and account for the repeated measurements, we used random slopes and intercepts (see below). For stroke participants, success was highly predictive of condition: success was 100% in the no-time constraint condition but largely reduced in the fast-time constraint condition. Thus, to reduce model complexity (that is, the number of parameters to estimate) and avoid high collinearity, we included success but not time constraint condition as a factor. In the control participant model, we only included effort, because success was near 100% for this group for both fast-time and no-time constraint conditions.

We developed the logistic regression models using a forward step-wise approach. We started with the simplest (base) model with mixed intercepts. Next, for the stroke participants, we added one predictor among effort, success, and paretic side (RH or LH). We then extended the model by including two predictors among effort, success, and paretic side, and finally with all three predictors as main effects, and with interactions between paretic side with effort and success.

For instance, for a RH stroke participant *j*, the model given the probability of using the more-affected arm for a target *i* for participant *j*, as a function of effort and success is given by the following equation:

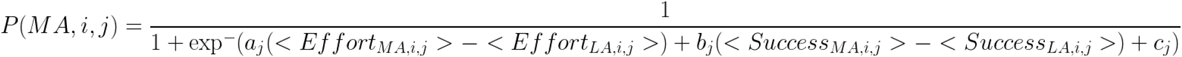

where, *C_j_* is a constant parameter that biases overall arm choice for the subject *j*, and *a_j_* and *b_j_* are weighting parameters for differences between the more-affected arm (MA) and less-affected arm (LA) in effort and success during the no-time and fast-time constraint conditions. The mixed-effect parameters for each participant *j* (*a_j_, b_j_*, and *c_j_*) are drawn from a Gaussian distribution centered on the mean of all participants (the fixed effects). The brackets < > indicate expected effort and success for movement to target *i* in subject *j*. Expected effort and success were computed from the movements to this target in forced choice blocks (see above). The probability of choosing the less-affected arm is given by *P_(LA,i,j)_* =1- *P_(MA,i,j_*_)_.

All predictors were z-transformed before inclusion in the models such that the parameters could be compared. We compared nested models using the Akaike Information Criteria (AIC) and with the Likelihood Ratio Tests (LRT) using the *anova()* function in *R*. Because of the data-driven forward approach model development and selection, we set a strict significance threshold of p = .005 for both model selection with the LRT and for significance of the fixed effects. Note that the models were developed with binary choice data from all participants in both stroke groups (stroke model, with choice data from Day 3) and control group (control model, with choice data from Day 2), with a maximum of 35 targets * 2 trials per target * 2 conditions = 140 data points for each participant. However, 4.5% of data for stroke group and 5.5% for control group were missing, yielding totals of 2936 and 1420 useful data points for the stroke and control models, respectively. Missing data was due to the 1) inability to obtain the whole reaching trajectory when participants used the opposite arm by mistakes in the forced choice block, or 2) abnormally long movement duration, as detected by > 3 standard deviations (such long durations were often due to unexpected movements such that the participants rubbed their eyes or noses).

### Switch - Stay analysis to determine the effect of failures on arm choices

We then assessed whether participants post-stroke switched to their less-affected arm in the spontaneous choice blocks when failing to reach targets with their more-affected arm in the forced choice blocks of the fast-time constraint condition. Specifically, for this analysis, we first considered the targets for which the participants used their more-affected arm in the spontaneous choice blocks in the no-time constraint condition. Next, among this first target sub-set, we selected the targets that were unsuccessfully reached with the more-affected arm in the forced choice blocks in the fast-time constraint condition. Finally, among this second target sub-set, we studied arm choice in the spontaneous choice blocks in the fast condition. The participants could either “switch” to the less-affected arm, or “stay” with the more-affected arm. Chi-square test was used to compare the proportion of subjects who switch and/or stay between the RH and LH groups

## Results

### Participant demographics and clinical data

Participants in the RH, LH, and control groups did not differ in age *(p* = .272, Kruskal-Wallis Test). The UEFM scores of participants ranged from 19 to 57 and were not different between the LH and RH groups (LH: 41.6 ± 3.3, RH: 44.9 ± 3.2, Mann-Whitney U test, *p* = .453). There was no difference in time since stroke onset between LH and RH groups (LH: 3.4 ± 1.5 years, RH: 2.2 ± .4 years, Mann-Whitney U test, *p* = .496). Consistent with the entry criteria, all participants reported to be right hand-dominant before stroke.

### Overall choice, success, and effort

Table 1 summarizes the overall probability of arm choice, success rate, and estimated effort of both arms for the RH, LH, and control groups in each condition. The RH group showed no change in more-affected arm’s choice across time constraint conditions *(p* = .840), whereas the LH group showed decreased in more-affected arm’s choice in the fast-time constraint condition compared to the no-time constraint condition *(p* < .001). The more-affected left arm choice in the LH group was lower than the more-affected right arm’s choice in the RH group in the fast time condition (p < .001), but not in the no time condition (p = .430) – See Table 1 and Figure 2A. The RH group showed decreased in right arm choice compared to the control group in both conditions (p<.001), but the LH group showed significant decreased in left arm choice compared to the control group only in the fast time condition (p<.001). Both stroke groups showed 100% success rates for the more-affected arm in the no-time condition (which is not surprising given that reaching to the farthest target within 5 seconds with the affected arm with an inclusion criterion), but their success rates significantly decreased in the fast-time constraint condition (t = 22.1*,p* < .001 and *t* = 28.4*,p* < .001 for the RH and LH groups, respectively). The success rates of the more-affected arm in the fast-time constraint condition did not differ across the two stroke groups, although there as a trend for greater success in the RH group (61.6 ± 1.4 vs 46.2 ± 1.4 for the RH and LH groups, respectively; *t* = 2.37, *p* = .110) (Table 1). Effort of the more-affected arm was also not different between the LH and RH groups, but effort in the fast-time constraint condition was higher than in the no-time constraint condition (t = 4.07, *p* < .001 for both groups).

**Table 1.**
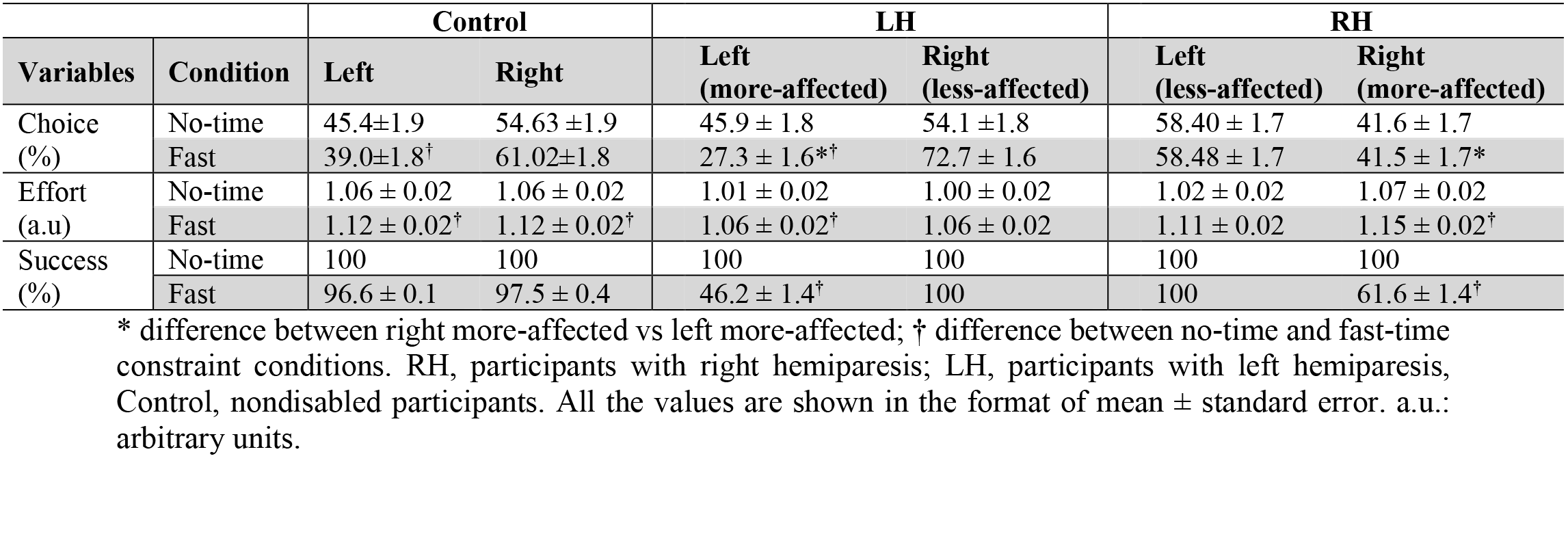
Choice, Effort, and Success for the right and the left arms across conditions in the RH, the LH, and the Control groups

**Figure 2.**
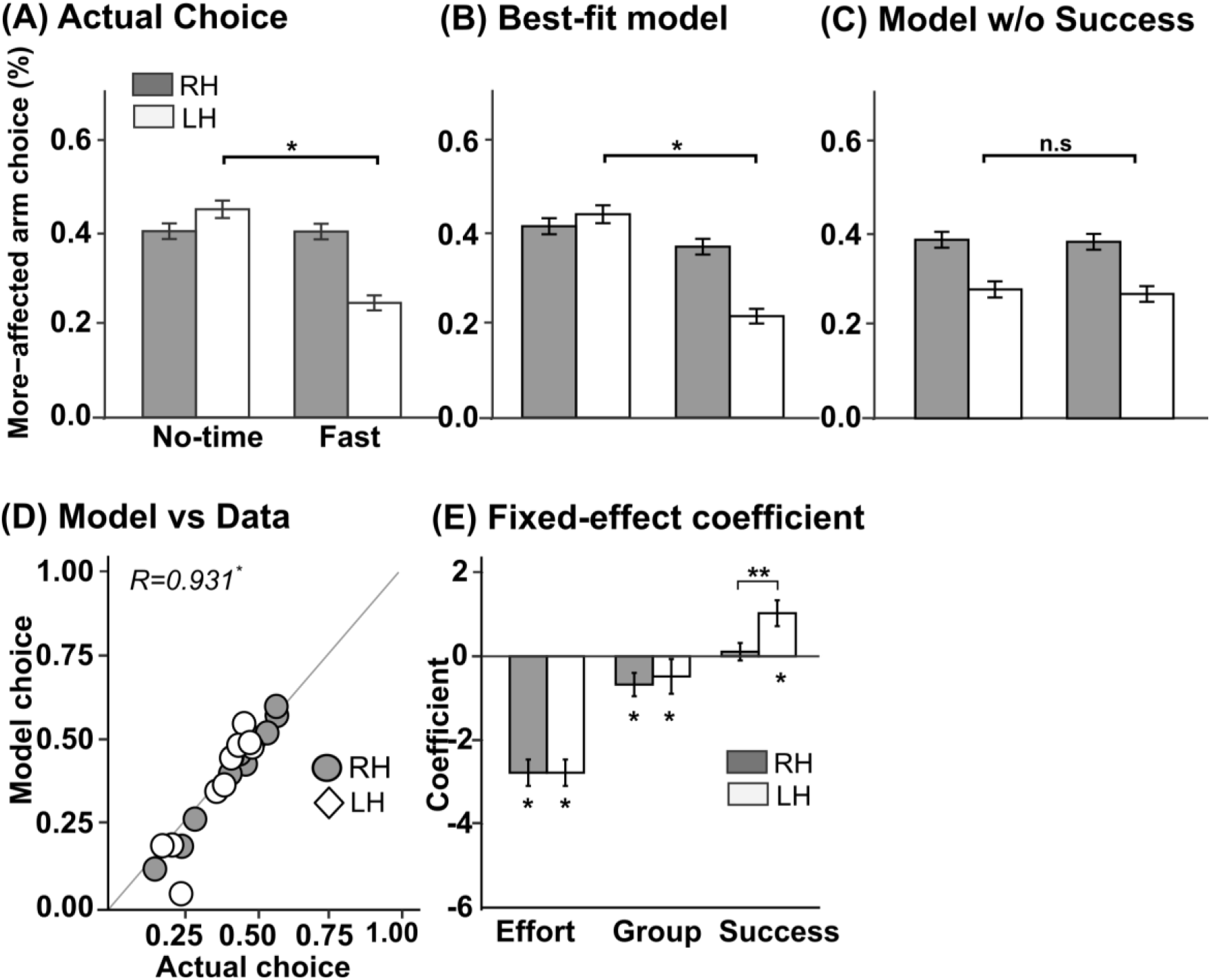
More-affected arm choices from actual experiment and results from best-fitted model. A. Actual choice for the RH (dark gray) and the LH (white) groups. B. The best-fit model predicts the more-affected arm choice accurately in both time conditions. C. A model without the success term predicts the more-affected arm choice less accurately for the LH group. The significant difference between the conditions in the LH group is indicated by *(p<.05) D. Average arm choice predicted by the model vs. actual choice in the fast condition for all participants post-stroke. E. Fixed-effect coefficients. The fixed-effect coefficient for success in the LH group is significantly lower than in the RH group, as noted by ** (p < .01). Fixed-effect coefficients significantly different from zero are indicated by * (p < .001).

### Expected success differentially influences arm choice in the RH and LH groups

We then tested whether the following regressors accounted for arm choice in individuals with stroke: 1) between-arm differences in effort, 2) between-arm differences in success, and 3) paretic side. For the stroke participants, Model 1 with effort showed a better fit (AIC = 2,330) than Model 2 with success (AIC = 3,648) and Model 3 with effort and paretic side (AIC = 2,332; all results are in Table 2). Model 4 with effort, paretic side, and interaction between effort and paretic side showed non-significant interactions (AIC = 2,332). The model with effort, paretic side, and success had a better model fit than Model 4 (Model 6; AIC = 2,147; comparison with Model 4; LRT, *p* < .001). Incorporating the interaction between success and paretic side without effort resulted in a poor model (Model 5). However, including the interaction between success and paretic side, together with effort resulted in the best fit (Model 7 AIC = 2,141; LRT: *p* = .003 between these two models). We thus selected Model 7 as our final model for arm choice in post-stroke.

**Table 2.** Mixed effect logistic regression models for stroke participants.

|  | Arm Choice Models |  |  |  |  |  |  |
| --- | --- | --- | --- | --- | --- | --- | --- |
|  | (1) | (2) | (3) | (4) | (5) | (6) | (7) |
| Constant(RH) | -0.789***<br>(0.181) | -0.539***<br>(0.119) | -0.692<br>(0.244) | -0.697<br>(0.244) | -0.389<br>(0.154) | -0.698<br>(0.280) | **-0.674**<br>**(0.279)** |
| Side(LH = 1) |  |  | -0.210<br>(0.359) | -0.196<br>(0.359) | -0.334<br>(0.230) | -0.404<br>(0.414) | **-0.480**<br>**(0.414)** |
| Effort | -2.610***<br>(0.328) |  | -2.608***<br>(0.327) | -2.964***<br>(0.424) |  | -2.783***<br>(0.315) | **-2.779\*\*\***<br>**(0.315)** |
| Success |  | 0.508***<br>(0.138) |  |  | 0.230<br>(0.159) | 0.566<br>(0.193) | **0.110**<br>**(0.209)** |
| Effort : Side |  |  |  | 0.797<br>(0.622) |  |  |  |
| Success : Side |  |  |  |  | 0.608<br>(0.236) |  | **1.029\*\*\***<br>**(0.309)** |
| Observations | 2,936 | 2,936 | 2,936 | 2,936 | 2,936 | 2,936 | **2,936** |
| Log Likelihood | -1,161.413 | -1,820.310 | -1,161.245 | -1,160.467 | -1,816.734 | -1,066.943 | **-1,062.568** |
| AIC | 2,330.827 | 3,648.621 | 2,332.490 | 2,332.935 | 3,645.468 | 2,147.886 | **2,141.136** |

AIC, Akaike Information Criterion; RH, participants with right hemiparesis; LH, participants with left hemiparesis. \*\*\* p < .005. The best-fit model is represented by bold.

For the control group, we performed a similar analysis and found that the effort term played a crucial role with no effect of condition (Table 3; note that success was not entered in this model because of the near 100% success rate for this group in all conditions, see Table 1).

**Table 3.** Mixed effect logistic regression models for the control participants.

|  | Arm Choice model |  |  |  |
| --- | --- | --- | --- | --- |
|  | (1) | (2) | (3) | (4) |
| Constant | **1.078\*\*\***<br>**(0.304)** | 0.187<br>(0.093) | 0.881<br>(0.313) | 0.916<br>(0.316) |
| Effort | **-4.131\*\*\***<br>**(1.054)** |  | -4.155***<br>(1.063) | -4.427***<br>(1.069) |
| Condition |  | 0.263<br>(0.108) | 0.401<br>(0.148) | 0.334<br>(0.151) |
| Effort:Condition |  |  |  | 0.498 |
| Observations | **1,420** | 1,420 | 1,420 | 1,420 |
| Log Likelihood | **-582.652** | -962.156 | -578.988 | -575.823 |
| Akaike Inf. Crit. | **1,173.303** | 1,930.313 | 1,167.976 | 1,163.646 |

AIC, Akaike Information Criterion; \*\*\*p < .005. The best-fit model is represented by bold.

Figure 2 shows overall predictions of the more-affected arm choice for all targets and all stroke participants from the selected Model 7. Comparison of average actual choice (Figure 2A) and average predicted choice (Figure 2B) indicates that the model captures the differences between the LH and RH groups and time constraint conditions, with a notable decrease in the more-affected arm’s choice in the fast condition for the LH group, but not for the RH group. Figure 2D, which shows the predicted vs. actual arm choice in the fast condition for the stroke participants, illustrates the excellent prediction of arm choice in the spontaneous choice blocks from data in the forced choice blocks *(Spearman’s correlation, r* = .93*,p* < .001). Accuracy of the model calculated by comparing the actual to predicted arm choice for each target, for the RH, LH, and control groups were 84.9 ±4.23%, 85.7 ±5.6%, and 82.8 ± 11.73 %, respectively.

The fixed-effect coefficient of effort was different from zero *(z* = -1.16, *p* < .001, see Model 7, Table 2) and there was no difference in this coefficient between the LH and RH groups (z = .88, *p* = .377) (Figure 2E). Whereas the fixed-effect coefficient of success was not different from zero in the RH group (z = 0.53*, p* = .600), it was greater than zero in the LH group (z = 3.33*,p* < .001). The importance of including success in the model is illustrated in Figure 2C, in which we compared arm choice in models that do and do not include the success term. Without the success term, predictions of arm choice largely deviate from the data in all time conditions for the LH group (Figure 2A and 2C). Together, these results show that success influences arm choice for the LH group, but not for the RH group.

Note that a possible confound in the above analysis is that, in the fast condition, the LH group exhibited a success rate with the more affected arm about 15% lower than that of the RH group. Thus, there was a possibility that higher success rates in RH diminished its influence in arm choice. We addressed this issue with two additional analyses. First, we matched participants in the two groups by success rates - this led to removal of two RH participants with the highest success rates (RH 1 & 6). Results did not change: the best fit model was still Model 7 and expected success still influenced arm choice in the LH but not in the RH groups (AIC for Model 6 = 2,012 and Model 7 = 2,006; LRT: *p* =.003 between these two models). Second, we fitted the models after removing one subject’s data from the overall data set, with each subject data removed in turn, leading to N = 22 model analyses. We verified that in these N = 22 analyses, the best-fit final model (as selected via the smallest AIC) was always Model 7.

### Switch - Stay differences between the RH and LH groups

We then analyzed individual trial data in both conditions across the no-time and fast-time constraint conditions to test whether participants maximized success by switching arms in spontaneous choice blocks when failing in the fast forced-choice condition with the more affected arm for the same targets (see Methods). Whereas the percentage of “stay” was not different between the RH and LH groups (p > .1), the percentage of “switch” was higher in the LH group than the RH group (56 ± 12% vs 22 ± 7%, p < .05; Figure 3A.). This indicates that the LH group switched to the less-affected arm more often when expecting unsuccessful trials in the fast condition than did the RH group. Thus, the LH group exhibited flexible behavior in arm choice to avoid unsuccessful trials, whereas the RH group appeared to be less sensitive to failures.

**Figure 3.**
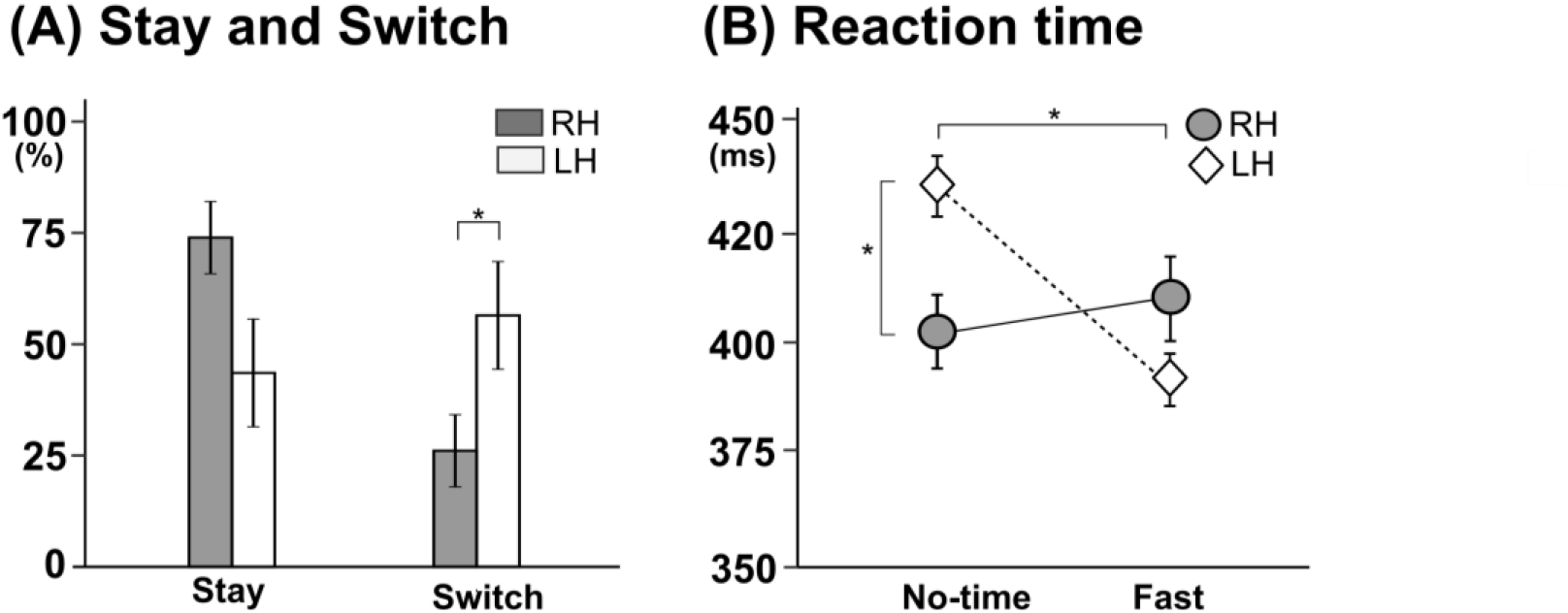
Stay and Switch and reaction times for the RH and the LH groups. A. Stay is defined as the percentage of trials in spontaneous choice block in fast time constraint condition in which subject keep on using their more-affected arm following failures in forced choice block in fast time constraint condition, and switch as the percentage of trials in which the subjects switched to the less-affected arm.. B. Reaction times measured in spontaneous choice block for both groups in no-time and fast time constraint conditions. The reaction times decreased from the no-time to the time constraint condition in the LH group, but not the RH group. RH, participants with right hemiparesis; LH, participants with left hemiparesis.* p<.05

### Reaction Times differences between the RH and LH groups

Figure 3B shows reaction times for groups and conditions. Reaction times of the spontaneous choice block in the LH group decreased from the no-time constraint condition to fast condition (no-time condition: 439 ± 7 ms and fast time: 390 ± 6 ms, *p* = .0012). However, the RH group showed no change in reaction times between the no-time and fast-time constraint condition (no-time: 403 ± 10 ms and fast time: 411 ± 11 ms, *p* = .598) (Figure 3B). In addition, the reaction times in the no time condition were larger in the LH group than in the RH group *(p* = .04).

**Figure 4.**
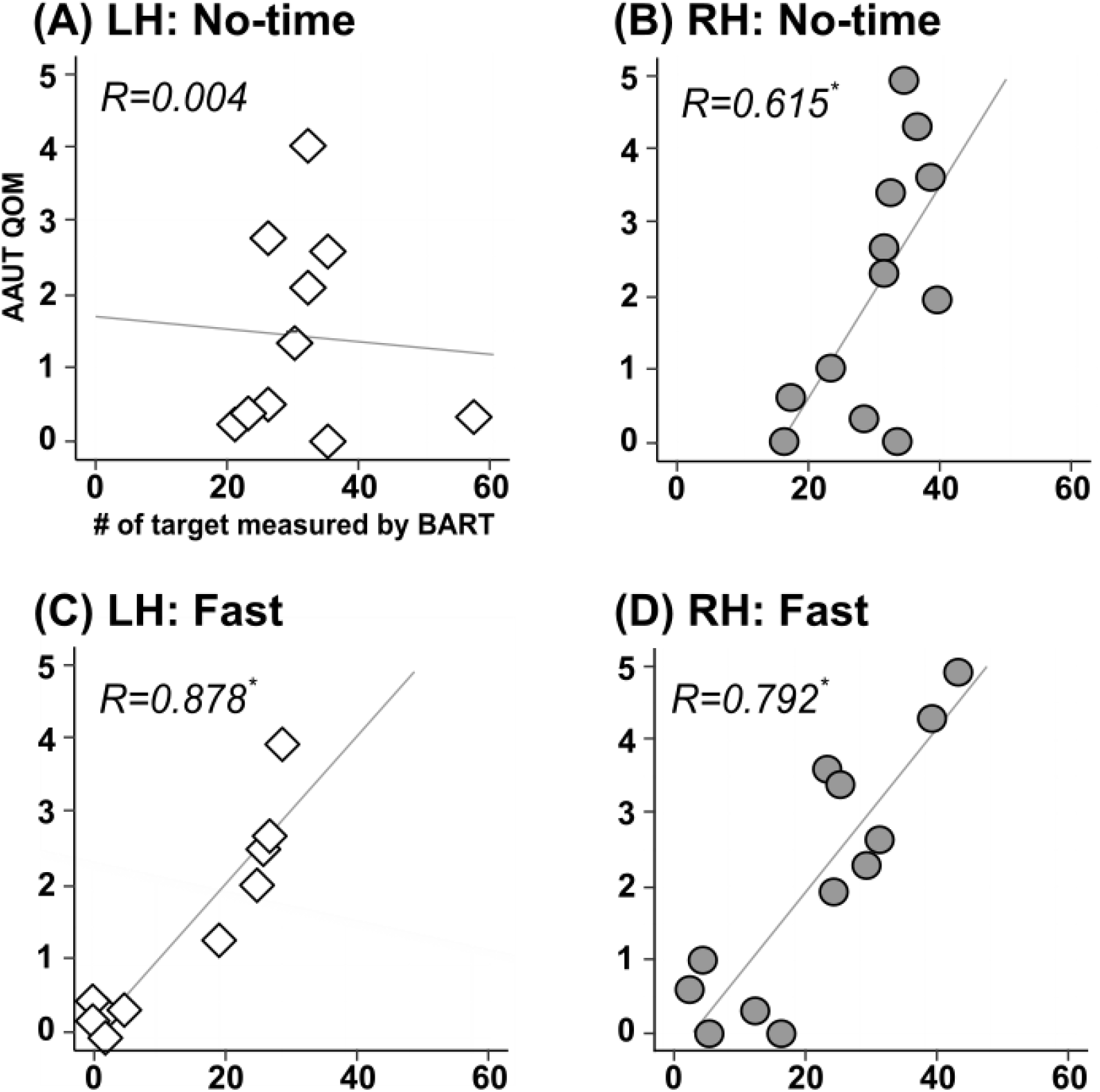
Correlations between AAUT and arm choice for RH and LH groups in no-time and fast time constraint conditions. Top row: correlations in no-time constraint condition for both RH and LH groups, Bottom row: correlations in fast time constraint conditions for both groups. The RH group shows good to moderate to good correlations between the AAUT score and arm choice under both conditions, while the LH group shows good correlation only in the fast constraint condition, but not in the no-time constraint condition.

### Clinical validation of arm choice with the AAUT

For the RH group, use of the more-affected arm measured by AAUT was significantly correlated with choice of the more-affected arm expressed by the number of targets that participants successfully reached using the more-affected arm under both the no-time and fast-time constraint conditions *(Pearson’s correlation, r* = .61, *p* =.023 for the no- time; *r* = .79, *p* < .001 for the fast-time condition- Figure 4 B,D). However, for the LH group, whereas arm choice of the more-affected arm in the fast condition correlated with AAUT *(Spearman’s correlation, r* = .88,*p* < .001), choice in the no-time constraint condition did not *(Spearman’s correlation, r* = .004,*p* = .91)(Figure 4 A,B).

## Discussion

We investigated how side of stroke, as well as effort and success, influence arm choice post-stroke. Our results overall support our hypotheses that in premorbidly right-handed individuals, RH individuals chose their paretic arm habitually, whereas LH individuals chose their paretic arm more optimally.

### RH individuals habitually use their more-affected arm, but LH individuals do not

Five complementary results from our study support greater habitual choice of the paretic arm in RH individuals than in LH individuals. First, whereas RH individuals showed no change in their overall probability of arm choice between the no-time and the fast-time constraint condition, LH participants showed a large decrease in choice in the fast-time constraint condition (see Table 1). Second, our logistic regression model of choice showed that the RH and LH groups differently responded to expected success: whereas the (fixed effect) parameter of expected success was near zero and non-significant in the RH group, it was significantly positive for the LH group. Removing the expected success term largely worsened the predictive capability of the model of choice for the LH group (Figure 2B and 2C). Third, compared to the LH group, participants in the RH group exhibited a small proportion of “switches” when given the choice of the arm to use in the fast condition, and thus persevered in choosing to perform unsuccessful reaching movements that often occurred after using the more-affected arm in this condition (Figure 3A). Fourth, reaction times of the RH participants did not change in the two time-constraint conditions (see Figure 3B), whereas reaction times of the LH participants largely increased in the no-time constraint condition compared to the fast-time constraint condition. Finally, a covert measure of arm use, the AAUT, correlated with arm choice for both the fast- and no-time constraint conditions in RH individuals (Figure 4). Because time pressure in the fast condition is thought to facilitate habitual choice (de Wit et al. 2018; Kim et al. 2018), via a shift from the goal-directed system to the habitual system (Keramati et al. 2011)(Hardwick et al. 2019), these results provide support for greater habitual choice of the paretic arm for RH individuals than for LH individuals.

Previous research on arm choice in both non-disabled young adults (Stoloff et al. 2011; Schweighofer et al. 2015) and in post-stroke individuals (Ballester et al. 2015, 2016) have shown that a competition between arms that results in decision to choose one arm is modulated by previous history of success. Such competition is thought to occur via neural populations in bilateral parietal cortices that encode actions in hand-specific terms and compete for action selection across and within hemispheres (Fitzpatrick et al. 2019). This competition has also been shown to be modulated by choices in the immediately preceding trials, leading to sub-optimal choices (Habagishi et al. 2014; Schweighofer et al. 2015; Valyear et al. 2019). Here, our results suggest that the long-term history of hand preference also modulates the competition between actions. Or results are consistent with the view that a long-lasting habitual bias, e.g. handedness or hand preference, modifies the competition between the two possible actions. Because habitual choices result in sub-optimal choices in RH participants, with fewer successes than could be achieved by choosing the less affected arm, why these long-lasting changes do not remap following negative experience? We can envision three possible non-exclusive possibilities that would counteract this remapping: First, the computational and time savings due to habitual actions in RH individuals could offset the advantages of otherwise more optimal decisions (Shadmehr and Krakauer 2008). Second, because the unsuccessful movement of the more-affected arm is often compensated with the less-affected arm in a bimanual manner in RH individuals (Haaland et al. 2012), remapping to avoid failures in daily activities might not be necessary. Third, repeated habitual choice in RH individuals can lead to (rewarding) improved function, in a form of self-training (Han et al. 2008; Hidaka et al. 2012), as suggested by the somewhat larger success rates in RH individuals compared to LH individuals despite overall similar impairment levels.

In contrast to the differential effect of expected success, the effect of the expected effort in arm choice was similar in RH and LH participants. Our results confirm, and extend to stroke survivors, the results of previous studies showing that nondisabled participants prefer to select targets, or the arm associated with lower biomechanical effort (Cos et al. 2011; Schweighofer et al. 2015; Wang et al. 2016; Bakker et al. 2017). Here, the effort parameter was the largest of all fixed parameters in all groups^1^; however, the parameter for effort was reduced in the post-stroke participants compared to the control participants. It is possible that, post-stroke, the computation of the expected effort, which requires a forward internal model of the dynamics, is not as accurate as in the healthy brain, leading to a relatively smaller contribution of effort to arm choice.

### Limitations and future work

The present study has several limitations and leaves several open questions that need to be addressed in future works. First, we estimated effort using a planar two-link arm (shoulder and elbow), although participants in our experiments could abduct their arm and rotate their shoulders internally or externally. In addition, our estimates of biomechanical effort do not take into account abnormal muscle co-contractions commonly shown in stroke survivors (Dewald et al. 1995; Ellis et al. 2005). Second, our experimental task may limit the generalization of our findings. In particular, both the use of the restraining belt to minimize compensatory trunk movements and the reaching movements that do not require finger movements or object manipulation may influence spontaneous choice. Third, it may be argued that our number of participants post-stroke (N=22) was too small to select the best model of arm choice and to test for the significance of three different regressors and two interaction terms. However, we believe that our results are valid because: 1) the stroke group was relatively homogenous, with strict entry criteria for the DOSE trial, 2) the models were pre-planned based on hypotheses derived from previous research, 3) the contributions, and relative sizes of the coefficients for the effort vs the success terms matched those of our previous study in non-disabled participants (Schweighofer et al. 2015), 4) we used a strict p = 0.005 for model selection and parameter significance, and 5) the stroke models were developed with multiple data points from all stroke participants with a maximum 140 arm choices per participants, which, accounting for missing data, yielded a totals of 2936 points. Fourth, because of the relatively small number of subjects, we could not link specific brain lesion characteristics to arm choice, nor could we include additional covariates in the models, such as impairment levels, pain, or the integrity of visuo-spatial memory, e.g., (Schweighofer et al. 2011). In future work with larger datasets, such covariates could be included to improve predictions of arm choice to a larger and more diverse population of stroke survivors.

### Implications of our findings for neurorehabilitation

Our findings, if confirmed with additional studies including reaching and grasping movements, suggest new rehabilitation strategies that are differently targeted to individuals with RH and LH. Spontaneous use of the more-affected arm in individuals post-stroke is a crucial indicator of both recovery and effectiveness of rehabilitation (Barker and Brauer 2005; Chen et al. 2012). Because stroke often leads to hemiparesis, individuals post-stroke often choose to perform motor actions with the less-affected arm. These individuals often exhibit at least some degree of non-use, that is, a decrease in use of the more-affected arm despite a residual capacity to use it (Andrews and Steward 1979; Taub et al. 1994; Hidaka et al. 2012; Han et al. 2013). Results from our arm choice model shows that for both RH and LH individuals, effort is a large (negative) predictor of arm choice. Thus, for all participants, treatment strategy should aim to increase spontaneous use by decreasing the more-affected arm’s effort through training which emphasizes fast movement and re-educates muscle co-activation patterns of shoulder and elbow to reduce abnormal elbow/shoulder joint torque coupling (Ellis et al. 2005, 2016). In addition, because success largely influence choice in the LH individuals, additional treatment regimens will be needed for these individuals to increase the use of their more-affected arm and to de-sensitize task failure in low-risk conditions (such as in this study) when the more-affected arm is used. Thus, for LH individuals, task oriented-repetitive movements with accompanying patient education, or virtual reality set-ups that encourage choice of the more-affected arm (Ballester et al. 2015, 2016), can be helpful to increase use in the natural environment (Winstein et al. 2016). In both cases, increased arm use may in turn further increase arm function via “self-training” (Hidaka et al. 2012); then the patient can enter a virtuous circle in which spontaneous arm use and motor performance reinforce each other (Han et al. 2008; Schweighofer et al. 2009).

## Data Availability

The data that support the findings are available on reasonable request.

## Acknowledgements

We thank members of the CNRL labs, Rebecca Lewthwaite, John Monterosso, and Jim Gordon for comments on this work. We thank Shannon Massimo and Nicolo Betoni for help with data collection and Hyeshin Park for help with BART programming.

* Author contributions: S.K. and N.S. wrote the paper; S.K., C.J.W., and N.S. designed the research; S.K. performed the experiments; S.K., C.E.H., B.K., and NS analyzed the data; C.J.W., B.K., and C.E.H. edited the paper.

* This work was funded by grants NIH R01 HD065438 and R56 NS100528.

* Conflict of interest statement: None

## Footnotes

1 As a reminder all variables were z-transformed so direct comparison of parameter values is meaningful.

